# Over-transmission of *NF1* mutant alleles in Neurofibromatosis type 1

**DOI:** 10.1101/2025.10.06.25337162

**Authors:** Yang Pei, Andrew Browne, Edgar Creus-Bachiller, Conxi Lázaro, Elisabeth Castellanos Perez, Meena Upadhyaya, Vincent M Riccardi, Margaret R Wallace, Anne Goriely

## Abstract

Neurofibromatosis type 1 (NF1) is one of the most common autosomal dominant tumor-predisposition syndromes (∼1:3,000 worldwide), caused by pathogenic variants in the *NF1* gene. NF1 is clinically diverse, involving pigmentary, skeletal, and neurodevelopmental features, alongside a lifelong risk of benign and malignant tumors. *NF1* encodes neurofibromin, a negative regulator of RAS-MAPK signaling, and behaves as a classic tumor suppressor, with tumorigenesis requiring biallelic inactivation.

Here, we analyzed transmission patterns in 322 NF1 families across four well-characterized cohorts, applying strict inclusion criteria to minimize ascertainment bias and avoid potential mosaic cases. Among 701 offspring, 61.1% were diagnosed with NF1, a significant excess over the 50% expected under Mendelian inheritance (p = 3 × 10⁻⁸). This transmission ratio distortion (TRD) was observed in both female (62.8%) and male (58.5%) transmitters. To test whether cohort size or other confounders could explain this bias, we performed sub-sampling and large-scale random down-sampling analyses, which confirmed robust TRD independent of parental sex or sample size.

We assessed plausible biological mechanisms for our findings and propose that the TRD observed in NF1 is most consistent with a novel mechanism involving clonal selection of *NF1*-null cells within the early embryonic germline, a concept rooted in established NF1 tumor biology. These findings uncover a novel aspect of NF1 biology with direct implications for clinical practice, prenatal diagnostics and reproductive counseling.

**Significance Statement:** Neurofibromatosis type 1 (NF1) is a common dominant cancer predisposition syndrome caused by mutations in the *NF1* gene. Using data from more than 700 offspring in 322 families, we show that mutant alleles are transmitted more frequently than expected, violating Mendelian inheritance. This phenomenon, known as transmission ratio distortion (TRD), is best explained by germline selection mechanisms analogous to those driving clonal expansion in NF1 tumors. Our findings reveal a previously unrecognized aspect of NF1 genetics with direct implications for reproductive counseling. More broadly, they suggest that germline selection may be an underrecognized driver of human inheritance and disease risk.

## Introduction

Although rare human disorders are inherently difficult to study, they often provide critical insights into fundamental biology. Among these, Neurofibromatosis type 1 (NF1, OMIM 162200), a well-characterized, autosomal dominantly inherited tumor predisposition syndrome affecting ∼1:3,000 individuals worldwide(1), has provided a wealth of new conceptual advances in genetics. NF1 has informed both tumorigenesis and neurodevelopmental biology, providing a rare conceptual link between cancer mechanisms and cognitive/behavioral disorders. It also exemplifies how tissue and developmental context influence the clinical manifestations of genetic mutations.

*NF1* is a large gene on chromosome 17q11.2, spanning ∼283 kilobases, with multiple isoforms encoding Neurofibromin (UNIPROT P21359), a 2839-residue protein that negatively regulates the RAS-MAPK signaling pathway. While clinical presentation is progressive and can be variable, even in families with the same mutation, heterozygous germline *NF1* mutations are typically associated with café-au-lait macules, skinfold freckling, and iris hamartomas (Lisch nodules). Additional features may include learning disability, autism or ADHD traits, skeletal abnormalities (macrocephaly, short stature, scoliosis, pseudoarthrosis), and tumors. Cutaneous peripheral nerve sheath tumors (neurofibromas) are almost universal in adults with *NF1*, and 10-15% of plexiform neurofibromas transform into malignant peripheral nerve sheath tumors (MPNSTs), a major cause of morbidity and mortality. At the cellular level, *NF1* acts as a classic tumor suppressor gene, with tumor formation following the “two-hit” model: heterozygous individuals requiring biallelic inactivation via loss-of-heterozygosity (LOH) or loss-of-function (LoF) mutations in specific cell types. Consistently, most neurofibromas harbor distinct, independent second-site *NF1* mutations (2, 3).

Another remarkable feature of NF1 biology, is its unusually high spontaneous germline mutation rate, reported to be significantly higher than that of most other disease genes, with some studies suggesting a 10-fold increase (4, 5). Consequently, ∼50% of *NF1* germline mutations occur sporadically as *de novo* mutations to unaffected couples. In addition, segmental NF1 caused by post-zygotic mosaicism is also commonly reported, while somatic *NF1* mutations are frequently found in tumors, contributing to tumor evolution, especially in leukemia, lung cancer and other solid tumors. A likely, but not often acknowledged, explanation for the high *NF1* spontaneous germline mutation rate is the contribution of selfish spermatogonial selection (6), a process by which spontaneously occurring mutations in the adult male germline are progressively enriched over time because they provide a selective advantage to mutant spermatogonial stem cells that outcompete their wild-type neighbors, thus increasing the proportion of mutant sperm with age. Studies of this process, that manifests in so-called Paternal Age Effect (PAE) disorders, have mainly implicated gain-of-function mutations in components of the RTK/RAS/MAPK pathway (6, 7), with best documented examples in *FGFR2* (i.e. Apert and Crouzon syndromes)*, FGFR3* (achondroplasia and thanatophoric dysplasia), *HRAS* (Costello syndrome) and *PTPN11* (Noonan syndrome). As a negative regulator of RAS/MAPK signaling, loss-of-function (LoF) mutations in *NF1* are anticipated to show the same effect and result in small clonal expansions in the testes of all men as they age (8, 9). However, whether selfish selection of *NF1* LoF mutations requires loss of one or both alleles in the testes of ageing men remains unclear. Note that since constitutional NF1 homozygosity (loss of function from both alleles) is considered an embryonic lethal condition, NF1 patients are assumed heterozygous for a germline mutation.

As an autosomal dominant disorder, NF1 has traditionally been thought to confer a 50% risk of transmitting the mutant allele to each offspring. Yet, reports have suggested the possibility of transmission ratio distortion (TRD) in NF1, whereby one allele is preferentially inherited by offspring of NF1 individuals, raising questions about potential deviations from Mendelian expectations. To date, these epidemiological studies have generally been small in scale and inconclusive, hindered by methodological and/or interpretive issues such as inconsistent adherence to the strict NF1 diagnostic criteria established in 1988 (10, 11), potential ascertainment bias in family-based studies (4, 12), and inclusion of *de novo* NF1 probands, some of whom may have been germline mosaic and therefore at reduced risk of transmitting the pathogenic variant.

Preimplantation genetic testing (PGT) offers an alternative means of assessing TRD, but previous results were inconsistent. Merker et al. (2015) analyzed 1060 embryo biopsies from 77 couples with one NF1-affected partner and observed a 54.4% transmission of the mutant allele, representing a significant deviation from the expected 50:50 ratio (two-sided exact binomial p = 0.00426) (12). In a similar approach, Vernimmen et al. (2023) analyzed 746 biopsied embryos from 82 couples in the Netherlands and found a 50.4% transmission rate, not significantly different from Mendelian expectation (13). However, only 30 couples (37%) in this latter cohort had familial NF1 mutations, while the majority were sporadic (56%) or presumed *de novo* cases (7%), again raising the possibility that parental germline mosaicism diluted the TRD strength.

Given the inconsistency of these reports and the unusual biology of NF1, we sought to assess the possibility of TRD by analyzing a large, combined cohort of NF1 families, applying strict criteria to minimize ascertainment bias. Here we report an analysis of 322 familial NF1 cases that revealed a statistically significant excess of affected offspring, consistent with transmission ratio distortion. To account for potential confounding factors, we applied different statistical approaches including stratification and down-sampling, which allowed us to systematically evaluate and exclude alternative explanations for the observed distortion. We then explore the most plausible biological mechanisms underlying the NF1 transmission distortion and consider its clinical implications for the counseling of NF1 families.

## Results

We collected a total of 322 families with demonstrable transmission of familial NF1 from a total of four study cohort groups from Spain (cohort S), the UK (cohort U) and the USA (cohorts V/W). Strict inclusion criteria were applied to avoid ascertainment bias and exclude potential mosaic cases (see Methods). Overall, eligible sibships included: (i) offspring of second-generation probands, (ii) siblings from prior generations other than the direct transmitting ancestor, (iii) offspring of affected aunts or uncles of the proband, and (iv) any subsequent generations from affected individuals in those sibships. Each sibship could therefore include no affected individuals, and only families in which the NF1 status was known for all offspring and both parents were included.

Counts for each cohort, stratified by the sex of the transmitting parent, are summarized in **Table 1** and **Figure 1A-B**. Maternal transmission occurred in 203 families (63%), and paternal transmission in 119 families. The aggregated cohort had a total of 701 offspring, corresponding to an average of 2.2 children per family (median = 2.0 children per family), with most families having 1-4 offsprings (**Figure 1B**). Male transmitters had a slightly larger number of offspring (*p* = 0.02539, unpaired t test), with an average of 2.4 children per family (median = 2.0), while female transmitters had an average of 2.1 children per family (median = 2.0).

**Figure 1.**
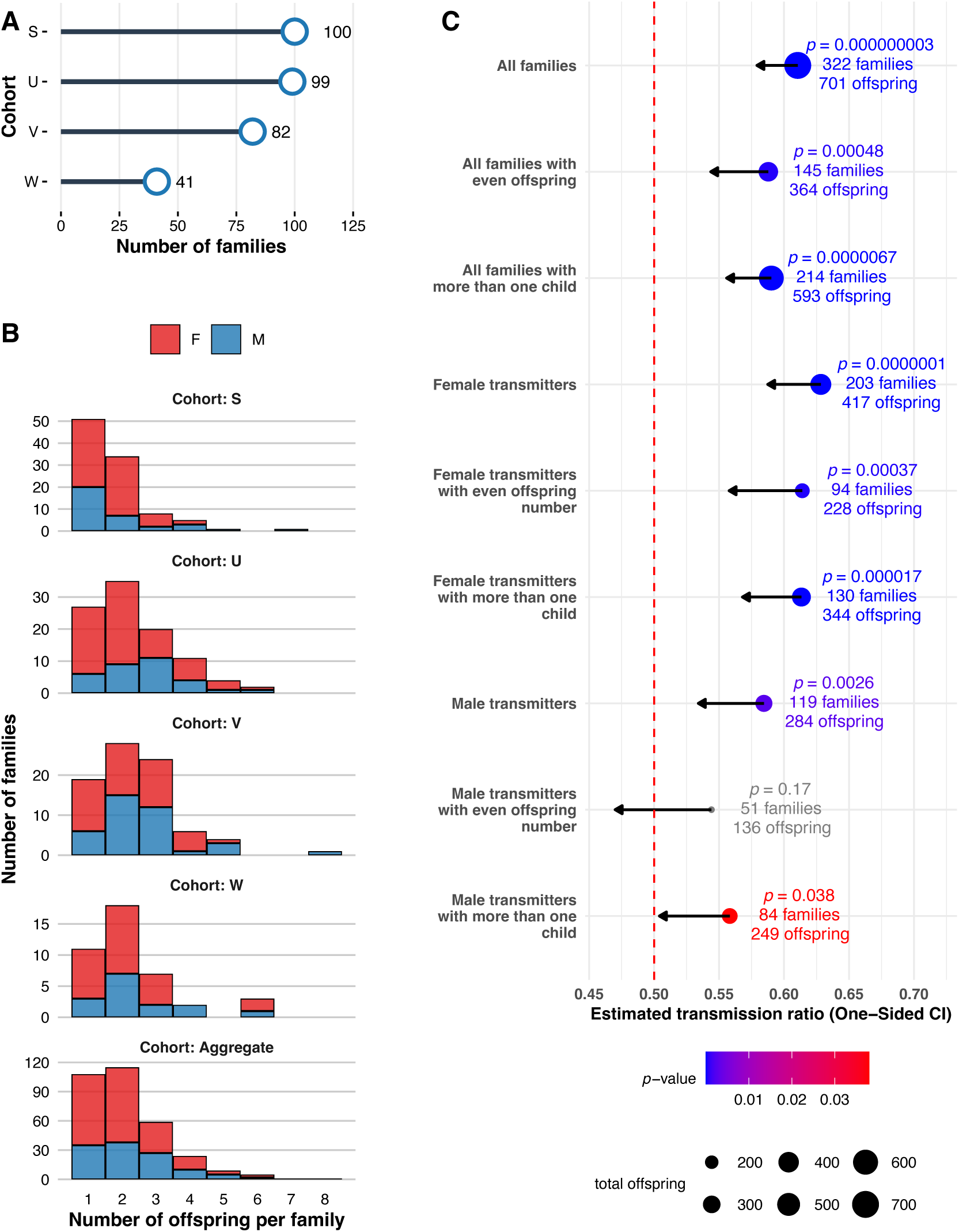
Characteristic features of the 322 NF1 families included in this study. *(A)* Number of families/sibships collected for the four NF1 cohorts (S = Spain; U = UK; V= USA1; W= USA2). *(B)* Distribution of family sizes per sibship count for each cohort and in aggregate (bottom), stratified by the sex of the transmitting parent: M=male transmitter (blue) and F=female transmitter (red). *(C)* Forecast plot illustrating the different data stratification approaches organized by estimated NF1 transmission ratio, including stratification by sex of transmitting parent to assess sex effects, excluding single-child families or families with an odd number of offspring to reduce extreme skews. P-value and lower confidence interval (arrow) were derived from one-tailed one-sample proportion test. The dotted red line represents the expected 50% random Mendelian segregation; size of the cohort and level of significance are described according to circle and color key, as indicated on the chart.

**Table 1.** Summary of the NF1 cohort characteristics and stratification strategies used to estimate NF1 TRD. (See Supplementary Table 1 for the complete dataset and statistical results)

| Condition | Families tested | Affected offspring | Total offspring | Transmission ratio | P Value* | CI low |
| --- | --- | --- | --- | --- | --- | --- |
| All families | 322 | 428 | 701 | 61.1% | 3.0E-09 | 57.9% |
| All families, even number of offspring | 145 | 214 | 364 | 58.8% | 4.8E-04 | 54.4% |
| All families, excluding single child family | 214 | 350 | 593 | 59.0% | 6.7E-06 | 55.6% |
| Female transmitter only | 203 | 262 | 417 | 62.8% | 1.0E-07 | 58.7% |
| Female transmitter only, even number of offspring | 94 | 140 | 228 | 61.4% | 3.7E-04 | 55.8% |
| Female transmitter only, excluding single child family | 130 | 211 | 344 | 61.3% | 1.7E-05 | 56.8% |
| Male transmitter only | 119 | 166 | 284 | 58.5% | 2.6E-03 | 53.4% |
| Male transmitter only, even number of offspring | 51 | 74 | 136 | 54.4% | 1.7E-01 | 47.0% |
| Male transmitter only, excluding single child family | 84 | 139 | 249 | 55.8% | 3.8E-02 | 50.4% |

To estimate the transmission ratio, we analyzed the transmission data from the aggregated cohort of 322 sibships (**Figure 1C**). Overall, 428 of 701 offspring were diagnosed with NF1 (61.1%), significantly exceeding the 50% expectation under Mendelian inheritance (one-sample proportion test, *p* = 3 × 10⁻⁸; **Figure 1C**; **Table 1**). To further investigate this unexpected result, we stratified the data by considering the sex of the transmitting parent. Increased transmission ratios above the expected 50% were observed in both male and female transmitter cohorts. Families with female transmitters (n = 203) showed a stronger bias, with 282 of 417 offspring affected (62.8%; one-sample proportion test*, p* = 1 × 10⁻⁷), whereas male transmitters (n = 119 families) produced 166 of 284 affected offspring (58.5%; p = 2.6 × 10⁻³). Although the transmission bias was more pronounced among female transmitters, the difference between maternal and paternal transmissions was not statistically significant (Chi-square test, χ² = 0.18, df = 1, *p* = 0.276). Additionally, among affected offspring, there was no significant sex bias (200 females vs. 212 males; two-sided binomial test with expected 50%, *p* = 0.588) (**Supplementary Table S1**).

Because the transmission ratios deviated from Mendelian expectation, we sub-sampled the cohort further to ensure that the results were robust and to rule out artifacts from sample heterogeneity or statistical noise. First, to avoid inflation of the signal by the large number of relatively small families, we repeated the analysis after excluding single-child families (n = 214 after removal of 108 single-child families). Single-child pedigrees tend to produce extreme transmission values that are not always informative and may bias the overall distribution toward the extremes. In this restricted dataset, non-single-child families showed a 59.0% transmission ratio (350 affected of 593 offspring across 214 families), a bias which remained significantly higher than the expected 50% (one-sample proportion test, *p* = 6.7 × 10⁻⁶; **Figure 1C**, **Table 1, Supplementary Table S2**). Hence, the significance of transmission distortion persisted even after removal of families prone to extreme values (i.e. single-child families).

Second, to address potential biases related to family structure, we excluded families with an odd number of offspring, as these families cannot produce an exact 50% transmission ratio under the null expectation of random segregation, and may therefore skew the findings. Analysis of the 145 families with an even number of offspring revealed a transmission ratio of 58.8%, which remained significantly higher than the expected 50% (one-sample proportion test, *p* = 4.8 × 10⁻⁴; **Figure 1C**, **Table 1, Supplementary Table S2**). This indicates that the inclusion of families with an odd number of offspring was not a major contributor to the observed transmission distortion.

Additionally, when stratifying by the sex of the transmitting parent in this restricted dataset of pedigrees with even number of offspring, sibships from male transmitters (n = 51; 136 offspring) showed a 54.4% transmission ratio, whereas families with female transmitters (n = 94; 262 offspring) showed a stronger bias at 61.4%. Only the female-transmitter group exhibited a transmission ratio significantly higher than the Mendelian expectation (*p* = 3.7 × 10⁻⁴), while the deviation in male-transmitter families was no longer statistically significant (*p* = 0.173). However, the ‘male-even’ sub-group was the smallest cohort (n = 51 families, 136 offspring), limiting statistical power and likely contributing to the lack of significance.

Because there were nearly twice as many female-transmitters as male-transmitters (203 vs. 119), we evaluated whether sample size differences contributed to the observed bias by randomly down-sampling the female-transmitter cohort to match the size of the male cohort. As shown in **Supplementary Figure S1**, the first 30 random down-sampling trials consistently produced higher transmission ratios in the female group than in the male-transmitter cohort, despite being matched for cohort size. Across all 500 down-sampling trials **(Supplementary Table S3)** only five yielded lower transmission in the female-transmitter cohort compared to the males. Although female-transmitters tended to show higher transmission rates overall, none of the individual size-matched female trials reached statistical significance (**Supplementary Figure S1; Supplementary Table S3**), consistent with the cohort-wide Chi-Square test result (*p*-value = 0.276).

Finally, to further assess the impact of sample size, we simulated datasets by randomly down-sampling the cohort across a range of family sizes. This analysis was designed to illustrate how limited sample size can misrepresent transmission distortion. Results were visualized by plotting the mean transmission ratio against the total number of offspring in each trial (**Figure 2, Supplementary Table S4**). As expected, smaller sample sizes produced greater variability in transmission ratios whereas larger sample sizes yielded more stable values. A consistent trend emerged: including more families, particularly those with larger sibship sizes, progressively narrowed the variance and converged on the transmission ratio observed in the full cohort. These results confirm that cohort size critically determines the stability of transmission ratio estimates, and that the 322 families (701 offspring) analyzed here provide sufficient power to yield robust and reliable estimates of transmission, and thus ability to detect TRD.

**Figure 2:**
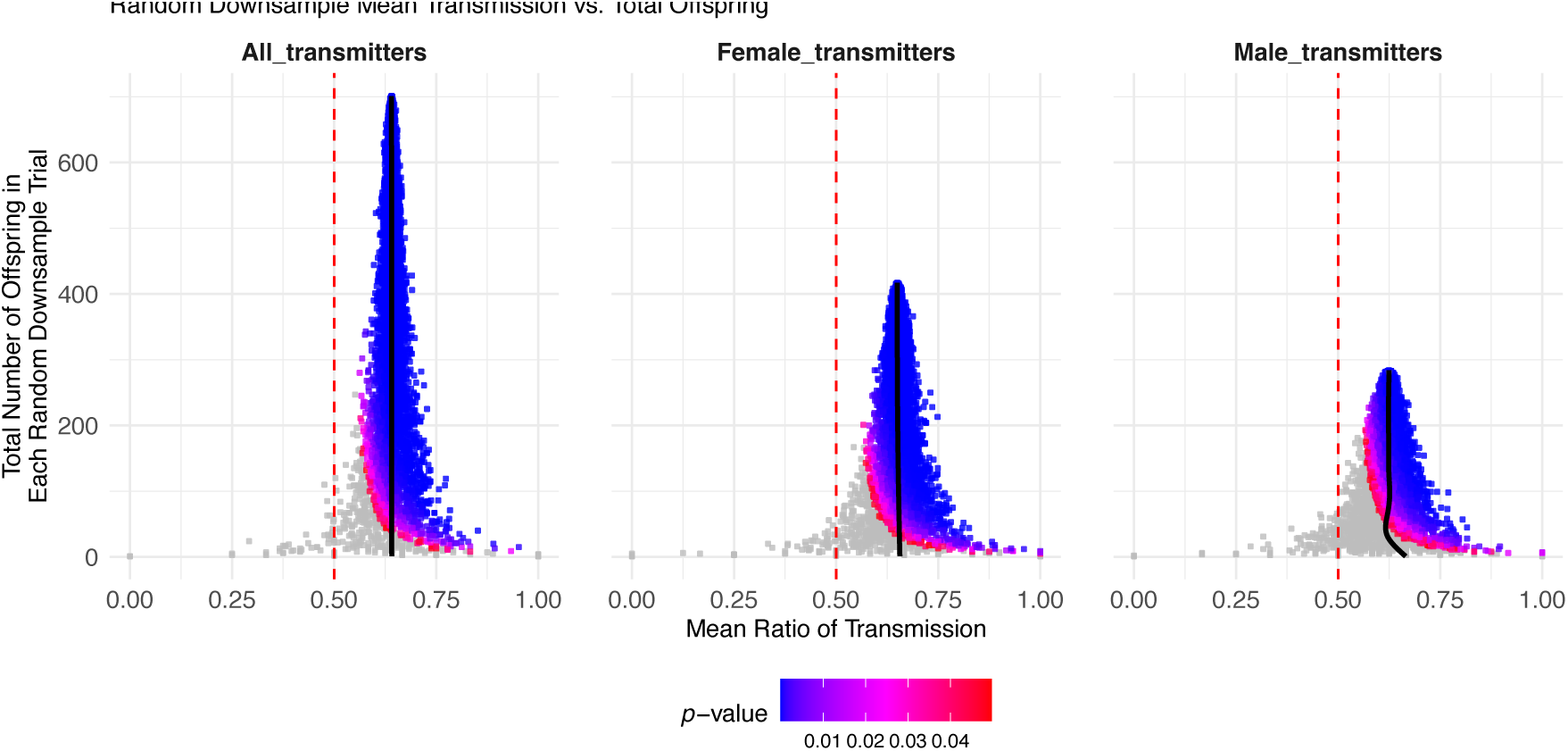
Illustration of the impact of sample size on transmission ratio: for the whole cohort (left) or split by sex of the transmitting parent (i.e. female transmitter (middle) or male transmitter (right). A total of 5,000 random down-sampling trials were performed at varying sample sizes to systematically assessed the effect of sample size on the estimated transmission ratio. For each trial, a one-tailed one-sample proportion test was conducted under the null hypothesis of 50% transmission ratio (dotted red line). The X-axis indicates the mean transmission ratio across all trials of a given sample size, rather than the observed transmission of the entire cohort as shown in Table 1. Each point represents one trial, plotted against the total number of offspring in that trial. Points are colored by p-value (see key on figure), with trials showing no statistical significance (p < 0.05) greyed out. Results of each individual trial are given in **Supplementary Table S3**.

## Discussion

In this study, we demonstrate that *NF1* mutations from heterozygous individuals are transmitted to offspring at significantly higher than expected rates, representing a clear deviation from Mendel’s second law of random segregation. Our analysis, based on data collected from 322 family sibships, each carefully ascertained and evaluated to minimize bias and control for potential confounders, showed a strong transmission ratio distortion (TRD), with 61.1% of offspring inheriting the mutant *NF1* allele.

Given the unexpected nature of this result, we systematically evaluated the impact of potential confounders, including the sex of the transmitting parent and the structure of the family pedigrees. We found that TRD was consistently observed under all stratification conditions. Importantly, although female transmitters exhibited slightly higher distortion (62.8% affected offspring) than male transmitters (58.8%), this difference was not statistically significant. We further accounted for possible artifacts from small sibship sizes, and the retrospective nature of clinical data by applying strict inclusion criteria, stratification analyses, down-sampling, and data simulations. Together, these approaches argue against major confounding bias, and the consistency of the findings across large sub-cohorts supports the conclusion that the observed TRD reflects a genuine biological phenomenon.

While segregation distortion is a well-recognized phenomenon in genetics, the biological mechanisms involved are not always fully understood (14). Notably, most evidence for TRD comes from studies in animal and plant models, with only few examples documented in human disease contexts (15-18). Below we briefly review mechanisms commonly implicated in TRD (14) and consider their potential relevance to our findings.

The best-characterized TRD mechanism is meiotic drive, in which ‘selfish’ alleles bias their own transmission through processes such as unequal chromosome segregation or selective gamete elimination. In females, driver alleles exploit the intrinsic asymmetry of meiosis to ensure they are retained in the oocyte, acting via altered centromere architecture, changes in kinetochore dynamics, or regulatory pathways involving aurora kinases (19, 20). In this context, the peri-centromeric location of the *NF1* gene may be relevant, and prior studies have implicated neurofibromin in chromosome segregation (21). By contrast, in males, meiotic drivers often act post-meiotically by sabotaging rival sperm. A classic example is the *t* haplotype in mice (22, 23) which impairs the motility of wild-type sperm, and thereby favors transmission of the driver allele. However, meiotic drive is an unlikely explanation for the NF1 TRD, since the effect was observed in both male and female transmitters. Most known drive systems are sex-specific, and invoking distinct male- and female-specific mechanisms seems unnecessarily complex (23, 24) and, although meiotic drive has been documented in a wide range of eukaryotic systems (25), no bona fide example has been conclusively described in humans.

Alternative explanations such as epigenetic factors (18), chromosomal abnormalities (26, 27) or mechanisms involving gametic (28) or zygotic (29) selection also appear unlikely to account for NF1 TRD. Notably, *NF1* is not an imprinted locus and shows no parent-of-origin effects, ruling out the involvement of allele-specific methylation. While chromosomal rearrangements can produce biased segregation, they occur too rarely to explain the strong and consistent distortion observed in our cohort. Gametic selection could, in principle, favor transmission from males if mutant sperm were more competitive, but this mechanism cannot account for the equally strong distortion seen in female transmitters. Finally, known examples of zygotic selection typically favor wild-type alleles and reduce the prevalence of pathogenic variants (29), a pattern opposite to the bias toward mutant *NF1* alleles identified in this cohort.

In contrast to the established TRD mechanisms above, we propose an alternative explanation, conceptually similar to the process of tumor development in NF1, involving bi-allelic loss of *NF1* and clonal selection of homozygous mutant cells (**Figure 3**). In this model, a recombination event or secondary deletion in embryonic germline tissues leads to *NF1* loss of heterozygosity (LOH). Cells that become functionally *NF1*-null through LOH, mutant homozygosity or compound heterozygosity gain a selective advantage in the *NF1* heterozygous background, allowing their clonal expansion within the developing gonad. During gametogenesis, these mosaic clones produce exclusively mutant haploid gametes, thereby driving the transmission ratio above the Mendelian expectation of 50%. This process is conceptually analogous to selfish spermatogonial selection documented for RTK/RAS/MAPK pathway mutations in the adult male germline (6, 7). However, to account for the sex-independent and pronounced distortion (∼61%) observed here, such mutational events would need to arise early in the developing germline lineage, at a developmental stage when only a small pool of primordial germ cells (PGCs) is present, and prior to sex determination. Supporting this scenario, *NF1* is expressed in both male and female embryonic (https://www.ebi.ac.uk/biostudies/arrayexpress/studies/E-MTAB-6592, last accessed Aug 2025) (30) and adult gonadal tissues (https://www.proteinatlas.org/ENSG00000196712-NF1/tissue https://www.gtexportal.org/home/gene/NF1, last accessed: Aug 2025) (31, 32).

**Figure 3:**
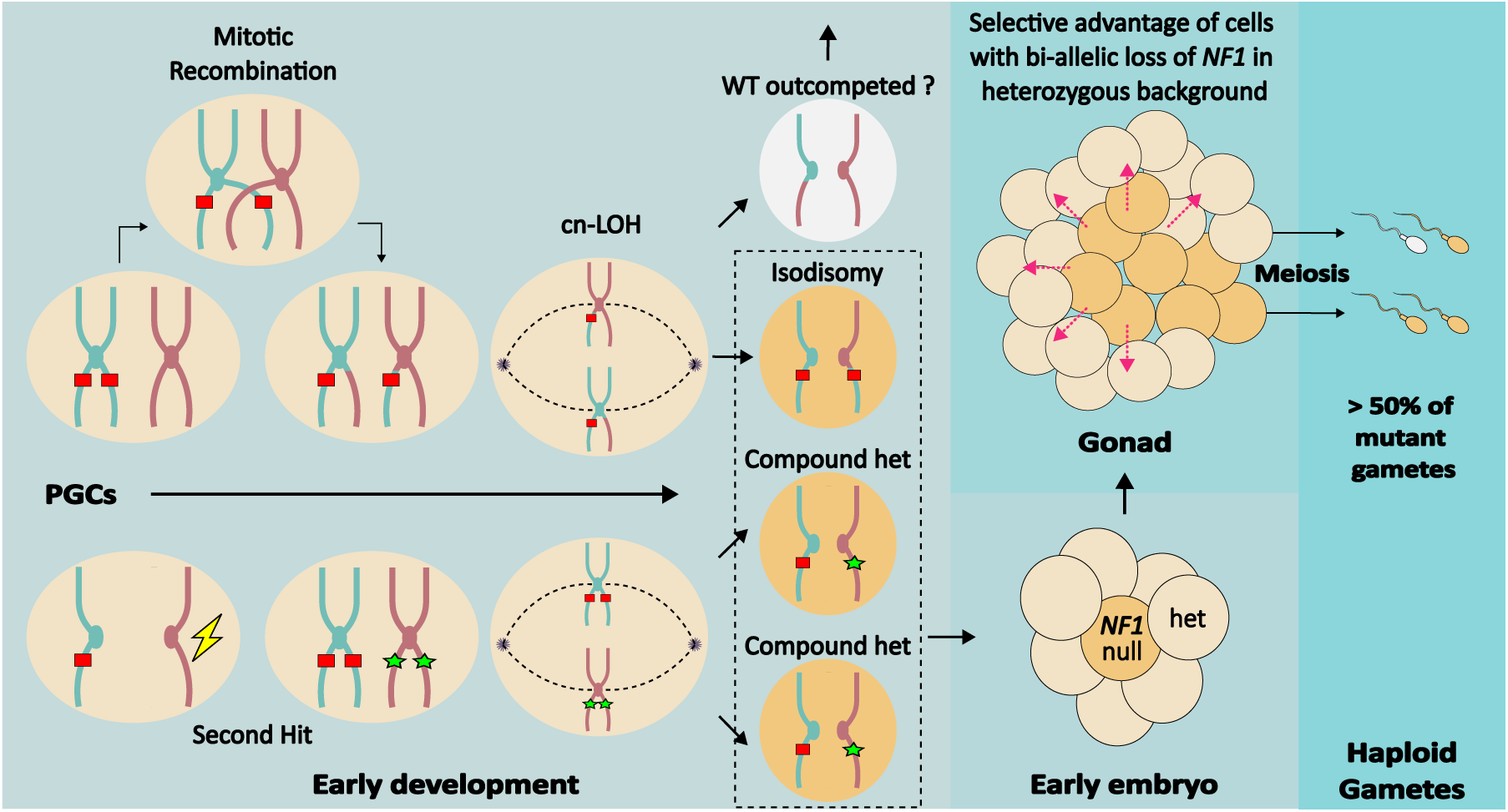
Proposed model for the TRD observed in familial NF1 cases. In this scenario, a random event of mitotic recombination between sister chromatids (top) or a second hit (i.e. point mutation or deletion) affecting the NF1-bearing chromosome 17 (yellow thunder) (i.e. compound heterozygosity) (bottom) within the developing germline tissue of a NF1 heterozygous individual can give rise to bi-allelic NF1 inactivation in the primordial germ cell (PGC) lineage. Bi-allelic loss of NF1 confers a selective advantage to null-mutant PGCs over their heterozygous counterparts, which clonally expands in the developing gonad (right). During gametogenesis, a higher-than-expected proportion of null-mutant haploid gametes (>50%) is produced, driven by the increased prevalence of NF1-null germ cells (oogonia or spermatogonia). To account for the observed sex-independent and pronounced transmission ratio distortion, such events must occur early in embryogenesis - at a developmental stage when only a small number of embryonic cells are present and before 6 weeks of development, the time at which sex determination occurs in humans. These mechanisms are analogous to that described in NF1-associated neoplasia, where NF1-null cells arise through cnLOH (at mitotic recombination hotspots), or as compound heterozygotes via deletion of the wild-type allele or following an acquired mutation on the second NF1 allele, i.e. a second hit (3, 41, 43). The red box and green star illustrate different mutations at the NF1 locus (i.e. deletion or point mutation) and the white, yellow or orange colours represent the NF1 wild-type (WT), heterozygous or null mutant status of the cell, respectively.

In this model, copy-neutral LOH (cnLOH), or uniparental disomy (UPD), are mechanisms of particular relevance. These result in homozygosity (i.e. isodisomy) without altering DNA copy number and is a well-documented driver of NF1 tumorigenesis (3). Due to rapid cell division and chromosome segregation errors, early embryogenesis is particularly vulnerable to cnLOH (33-35). When such a cnLOH event occurs within the developing germline lineage, it generates a subpopulation of homozygous *NF1*-mutant gamete-producing cells. Importantly, mitotic recombination at well-defined hotspots in and around *NF1* is a known pathogenic mechanism in NF1-associated tumors (36-38), being observed in ∼50% of neurofibromas (39-42) and even reported in normal CNS-derived tissue in a young NF1 patient (43).

Hence, a parsimonious model emerges in which positive selection of *NF1-*null mutant cells occurs in the developing germline lineage. This explanation is attractive because it builds on a well-established somatic process in NF1 tumorigenesis and naturally accounts for the consistent transmission bias observed in both male and female transmitters in our cohort.

Given that NF1 is a clinically significant genetic target, with many patients, often from large families, our findings have important implications for clinical practice and genetic counseling, including prenatal and preimplantation testing, which are often sought because of the unpredictable phenotypic impact of *NF1* mutations. Our finding that transmission risk exceeds 50% suggests that current reproductive guidelines may require updating. This elevated risk, combined with the NF1 unpredictable severity, could significantly influence family planning of individual families.

Another important consideration for such NF1 testing– i.e. prenatal diagnostics, via chorionic villus sampling, amniocentesis, or non-invasive cell-free fetal DNA analysis (44) – is that interpretation would be impacted by whether the TDR arose from cnLOH (via mitotic recombination/UPD), compound heterozygosity (new mutation on the second allele (**Figure 3**)) (3, 41, 43), or yet another process. This distinction is critical for prenatal diagnostics: if a new mutation on the second allele is involved, routine haplotype-based testing may fail to detect it. Because the NF1 gene is large and highly heterogeneous (>5,000 pathogenic variants reported in ClinVar https://www.ncbi.nlm.nih.gov/clinvar), most familial testing involves haplotype-based linkage analysis using microsatellite markers. While this allows prenatal diagnosis through detection of the parental risk haplotype, under the “second hit” hypothesis such testing may fail to detect the new *NF1* variant. Importantly, independent *NF1* mutations within the same family, although infrequent, have been documented in several reports (45-49).

Elucidating the mechanisms underlying TRD could most feasibly be achieved through sperm analysis in NF1 males, for whom gamete sampling is straightforward (50).

Quantifying the proportion of sperm carrying the familial *NF1* variant and screening for other *NF1* mutations by deep-sequencing of the full gene would also provide individualized transmission risk assessments prior to pregnancy (50).

As highlighted by our sample-size simulations, larger scale studies with rigorous diagnostic and inclusion criteria are needed to further validate and extend our findings. Incorporating *NF1* mutation data and expanding the analysis to families from diverse ethnic backgrounds will increase statistical power and may reveal whether specific allele types are more prone to TRD. A critical next step will be the direct analysis of sperm from NF1 males to assess both the frequency and underlying mechanism of TRD (i.e., mosaic mitotic recombination or second-hit events). Such studies will also refine reproductive counseling guidelines, and enable personalized interventions to support NF1 families in reproductive decision-making (50).

In conclusion, by documenting TRD, our study reveals a previously unrecognized facet of the NF1 genetics. We propose that NF1’s well-established somatic pathogenic mechanism also operates in the germline. This model provides a parsimonious explanation for the TRD observed in our large cohort. Notably, TDRs have also been reported in other cancer predisposition syndromes, including Retinoblastoma (*RB1*) (17) and Li-Fraumeni syndrome (*TP53*) (15), suggesting this may represent a broader phenomenon in human disease than currently recognized.

## Materials and Methods

Collection of NF1 transmission data from archival clinical pedigrees was performed following strict criteria to ensure that only familial transmissions from NF1 heterozygous individuals were considered. A ‘family’ unit or sibship was defined as two parents and their offspring, with the transmitting parent being an affected child of an NF1 parent (i.e., second or later generation cases. Families in which both parents had NF1 were excluded, and only multi-generation pedigrees were included. In some pedigrees, longitudinal data allowed inclusion of additional sibships from lower branches. Families were included only if NF1 status of parents and all siblings unequivocally met clinical diagnostic criteria, in some cases confirmed by molecular testing (51). Further, to minimize ascertainment bias and avoid over-representation of cases, the following approaches were used: (1) pedigrees specifically ascertained for linkage or pedigree studies were excluded, except for sibships in branches other than that of the study’s proband (i.e. proband’s generation and below); (2) the proband’s own sibship was excluded and (3) when counting sibships in prior generations, the transmitting parent was not included. Only sibships in which the affected parent was a familial case (i.e., also had an affected parent) were retained.

This study used existing de-identified NF1 pedigree data, under Exempt status (University of Florida IRB approval). In total, we obtained pedigree data from independent cohorts previously collected by four groups for clinical or non-linkage research purposes over ∼40 years (denoted as S [Spain], and U [UK], V [USA] and W [USA]). We analyzed transmission of NF1 from each affected parent to their offspring. For each family unit (referred to as a sibship), five primary data records were collected: (i) a unique family ID and the proband (first ascertained case), (ii) sex of the transmitting parent, (iii) number of affected offspring per sibship, (iv) total number of offspring per sibship and (v) number of affected female offspring per sibship. Three additional data points were derived from these records, as follows: (6) number of affected male offspring = number of affected offspring per sibship − number of affected female offspring per sibship; (7) number of unaffected offspring per sibship = total number of offspring per sibship − number of affected offspring per sibship; and (8) Transmission ratio (mutant) = number of affected offspring per sibship / total number of offspring per sibship (**Supplementary Table S1**).

The estimated transmission rate is the pooled transmission ratio derived as follow: All analyses were conducted using R Statistical Software (v4.4.1; R Core Team, 2024).

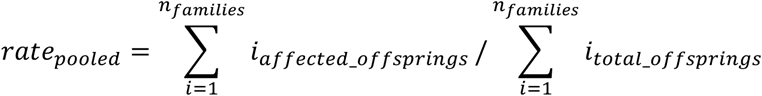

Given an expected transmission rate of 50%, a one-sided proportion test (using the prop.test function from R stats v4.4.1) was performed with the null hypothesis that the transmission rate is ≤50%. This test was applied to the aggregated cohort as well as seven sub-cohorts (**Supplementary Table S2**) including:

1. Cohort excluding single-child families.
2. Male-transmitter families.
3. Female-transmitter families.
4. Male-transmitter families with an even number of offspring.
5. Female-transmitter families with an even number of offspring.
6. Male-transmitter families excluding single-child families.
7. Female-transmitter families excluding single-child families.

To evaluate the impact of cohort size on male- and female-transmitter cohorts, we performed random down-sampling followed by a one-sided proportion test (**Supplementary Table S3**). Specifically, 500 random subsets of the female-transmitter cohort were down-sampled were generated to match the size of male-transmitter cohort (119 families).

Lastly, random down-sampling was performed at varying sample sizes for 5,000 trials for the whole cohort, as well as separately for the male- and female-transmitter sub-cohorts (**Supplementary Table S4**). Specifically, the respective cohort was randomly sampled with size ranging from 1 to 322 (all families), 1 to 203 (female-transmitters), and 1 to 119 (male-transmitters) in each trial. A one-sided one sample proportion test was carried out in each down sample trial to determine if the proportion of affected offspring was significantly greater than the expected 50%. The results were visualized by plotting the cohort size, i.e. total number of offsprings, from each trial against the corresponding estimated transmission ratio. Note that the observed transmission rate is the mean of individual familial transmission ratios derived as follow:

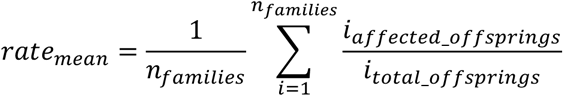

## Data Availability

All data produced in the present work are contained in the manuscript

## Acknowledgments

The authors wish to thank Dr Sue Huson, Alan Fryer, and the late Prof. Peter Harper for clinical assessment of UK families; all the NF1 families and patient associations for their support.

This work was primarily supported by grants from the Wellcome (219476/Z/19/Z) and the National Institute for Health Research (NIHR) Oxford Biomedical Research Centre Programme.

This Spanish team acknowledges the support of the Carlos III National Health Institute (PI23/00017, PI19/00553, PMP22/00064 and CPP2022-009550), the Centre de Recerca de Catalunya (CERCA) Programme (2021SGR01112), the European Commission (NextGenerationEU), the Fundación Proyeco Neurofibromatosis (FPNF) and the Fundació La Marató de TV3.

The funders had no role in study design, data collection and analysis, decision to publish, or preparation of the manuscript.

## Author Contributions

Conceived and supervised the study: MRW, VMR and AG Provided patient cohort data: CL, EC-B, ECP, MRW, MU, VMR Performed the data analysis: YP, AB, MRW and AG Wrote the manuscript: YP and AG with input from MRW and MU. All authors read and approved the final version.

## Competing Interest Statement

The authors declare no competing interests

This manuscript is dedicated to Vic Riccardi (1940-2024) in recognition of his contribution to the neurofibromatosis community over the last 50 years.

