## Supplemental Figure 1 for "Over-transmission of *NF1* mutant alleles in Neurofibromatosis type 1"

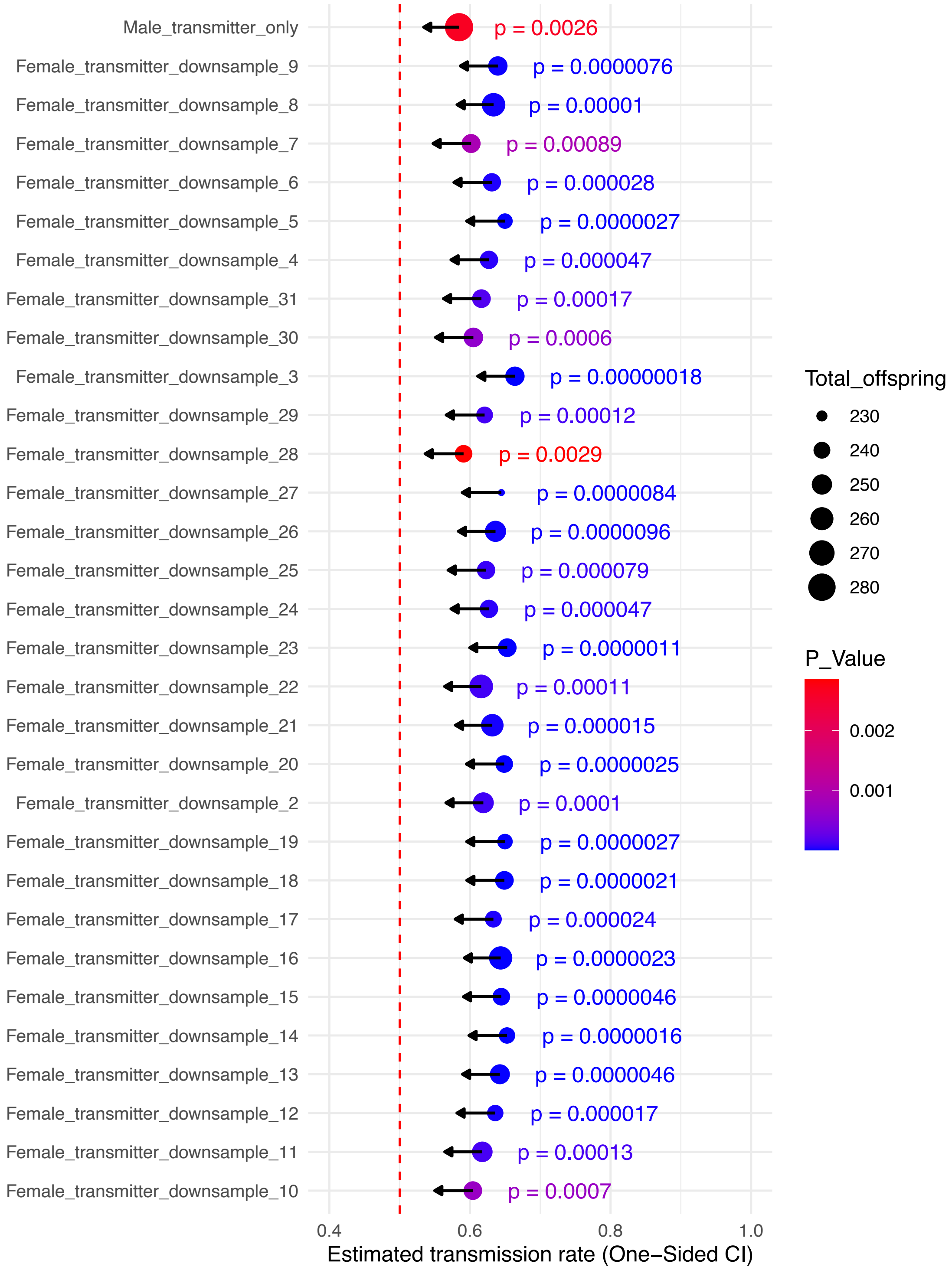


**Fig. S1.** Illustration of downsampling trials for the female transmitter dataset, aiming to match the female cohort size to that of the male cohort to assess whether the higher transmission ratio distortion observed in females can be explained by sample size. The first 30 random trials are shown, each with a corresponding one-tailed proportion test.
